# Disease overarching mechanisms that explain and predict outcome of patients with high cardiovascular risk: rationale and design of the Berlin Long-term Observation of Vascular Events (BeLOVE) study

**DOI:** 10.1101/19001024

**Authors:** Bob Siegerink, Joachim Weber, Michael Ahmadi, Kai-Uwe Eckardt, Frank Edelmann, Matthias Endres, Holger Gerhardt, Kathrin Haubold, Norbert Hübner, Ulf Landmesser, David Leistner, Knut Mai, Dominik N. Müller, Burkert Pieske, Geraldine Rauch, Sein Schmidt, Kai Schmidt-Ott, Jeanette Schulz-Menger, Joachim Spranger, Tobias Pischon

**Author notes:** Correspondence to Tobias Pischon. Equal contributions (Bob Siegerink, Joachim Weber).

## Abstract

**Background:** Cardiovascular disease (CVD) is the leading cause of premature death worldwide. Effective and individualized treatment requires exact knowledge about both risk factors and risk estimation. Most evidence for risk prediction currently comes from population-based studies on first incident cardiovascular events. In contrast, little is known about the relevance of risk factors for the outcome of patients with established CVD or those who are at high risk of CVD, including patients with type 2 diabetes. In addition, most studies focus on individual diseases, whereas less is known about disease overarching risk factors and cross-over risk.

**Aim:** The aim of BeLOVE is to improve short- and long-term prediction and mechanistic understanding of cardiovascular disease progression and outcomes in very high-risk patients, both in the acute as well as in the chronic phase, in order to provide the basis for improved, individualized management.

**Study design:** BeLOVE is an observational prospective cohort study of patients of both sexes aged >18 in selected Berlin hospitals, who have a high risk of future cardiovascular events, including patients with a history of acute coronary syndrome (ACS), acute stroke (AS), acute heart failure (AHF), acute kidney injury (AKI) or type 2 diabetes with manifest target-organ damage. BeLOVE includes 2 subcohorts: The acute subcohort includes 6500 patients with ACS, AS, AHF, or AKI within 2-8 days after their qualifying event, who undergo a structured interview about medical history as well as blood sample collection. The chronic subcohort includes 6000 patients with ACS, AS, AHF, or AKI 90 days after event, and patients with type 2 diabetes (T2DM) and target-organ damage. These patients undergo a 6-8 hour deep phenotyping program, including detailed clinical phenotyping from a cardiological, neurological and metabolic perspective, questionnaires including patient-reported outcome measures (PROMs)as well as magnetic resonance imaging. Several biological samples are collected (i.e. blood, urine, saliva, stool) with blood samples collected in a fasting state, as well as after a metabolic challenge (either nutritional or cardiopulmonary exercise stress test). Ascertainment of major adverse cardiovascular events (MACE) will be performed in all patients using a combination of active and passive follow-up procedures, such as on-site visits (if applicable), telephone interviews, review of medical charts, and links to local health authorities. Additional phenotyping visits are planned at 2, 5 and 10 years after inclusion into the chronic subcohort.

**Future perspective:** BeLOVE provides a unique opportunity to study both the short- and long-term disease course of patients at high cardiovascular risk through innovative and extensive deep phenotyping. Moreover, the unique study design provides opportunities for acute and post-acute inclusion and allows us to derive two non-nested yet overlapping sub-cohorts, tailored for upcoming research questions. Thereby, we aim to study disease-overarching research questions, to understand crossover risk, and to find similarities and differences between clinical phenotypes of patients at high cardiovascular risk.

## Introduction

Cardiovascular disease (CVD) is the leading cause of premature death worldwide and also the most common cause of permanent disability [1,2]. It is associated with organ dysfunction, multiple sequelae, impairment of quality of life as well as enhanced frailty and dependency in the elderly. Effective and individualized treatment requires exact knowledge about both risk factors and risk estimation. Most evidence for risk prediction currently comes from population-based studies on the risk of first incident cardiovascular events, such as the Framingham Heart Study[3], or the SCORE project[4]. In contrast, less is known about patients with established CVD or those who are at high risk of CVD, including patients with type 2 diabetes and target-organ damage. Currently, guidelines classify these patients as “high risk”, and assume that they have a >20% 10-year risk of recurrent events [5]. However, recent studies suggest that risk among these patients may vary substantially from <10% to >30%[6].

Cardiovascular diseases, such as acute coronary syndrome (ACS), acute heart failure (AHF) acute kidney injury (AKI), or acute stroke (AS) share many risk factors [7], including hypertension, lipid disorders, obesity, smoking, and others. Type 2 diabetes is also a major cardiovascular risk factor, and, when combined with target-organ damage, considered a high risk condition equivalent to manifest CVD [8–11]. Survivors of cardiovascular events often experience recurrences of the same disease and have largely increased risks for other cardiovascular events (crossover-risk) [12–17]. Not surprisingly then, an optimized therapy for one CVD condition may also reduce the risk for all vascular events and disease manifestations[18,19]. Still, despite these similarities e.g. cerebro- and cardiovascular diseases often manifest with different incidence rates, mortality rates, and recurrences [15,20,21]. Further, significant differences have been noted in the importance of predisposing risk factors for developing a certain vascular disease [22–24]. For example, smoking and LDL-cholesterol levels are more strongly related to the risk of ACS as compared to AS, whereas hypertension is a stronger risk factor in AS than in ACS. Substantial differences also exist between the pathomechanisms underlying these diseases [25]; e.g. atherosclerosis reveals considerable disparities between carotid and coronary artery disease [26]. Although the assessment [27] and prevention [1] of CVD risk factors such as hypertension, diabetes mellitus, and smoking have long been studied, preventive strategies often remain controversial [28–30].

More personalized risk stratification is necessary, which, however, requires a better understanding of the role of different risk factors in individuals. Deep phenotyping, where data is collected on the patient health status far beyond traditional risk factors, is likely to add a new dimension to subgroup identification and risk stratification. For example, systemic conditions such as immunological processes, [31] metabolic parameters [32] or the gut microbiome [33]. are largely understudied. Although in recent years deep phenotyping efforts have been part of larger scale population-based cohort studies, this is less so in the field of secondary prevention, and it is usually restricted to specific disease cohorts, whereas there is a paucity of studies that investigate cardiovascular disease overarching risk factors despite clear connections [34,35].

Therefore, we have initiated the Berlin Long-term Observation of Vascular Events(BeLOVE) study, a prospective deep-phenotyping cohort study which includes patients with five distinct disease phenotypes, to study and understand disease overarching risk factors and pathophysiological mechanisms which will help improve the risk prediction of morbidity and mortality.

### The overall aim of BeLOVE

The aim of BeLOVE is to improve short- and long-term prediction and mechanistic understanding of cardiovascular disease progression and risk in high-risk patients, both in the acute as well as the chronic phase. The objective of BeLOVE is to provide the basis for improved, individualized risk management in the long and short term.

### Study design

BeLOVE is a prospective cohort study of patients with a high risk of future cardiovascular events, including patients with a history of acute coronary syndrome (ACS), acute stroke (AS), acute heart failure (AHF), or acute kidney injury (AKI) or patients with type 2 diabetes with target-organ damage (T2DM). BeLOVE encompasses two partly overlapping subcohorts (Figure 1). The acute subcohort (subcohort 1) will recruite up to 6500 patients with ACS, AS, AHF, or AKI up to 8 days after their qualifying event. They will undergo a short structured interview on their demographics and medical history, a short cognitive test as well as quality of life measures, blood sampling. Patients will then be followed long term for up to 10 years by annual telephone interviews.

**Figure 1:**
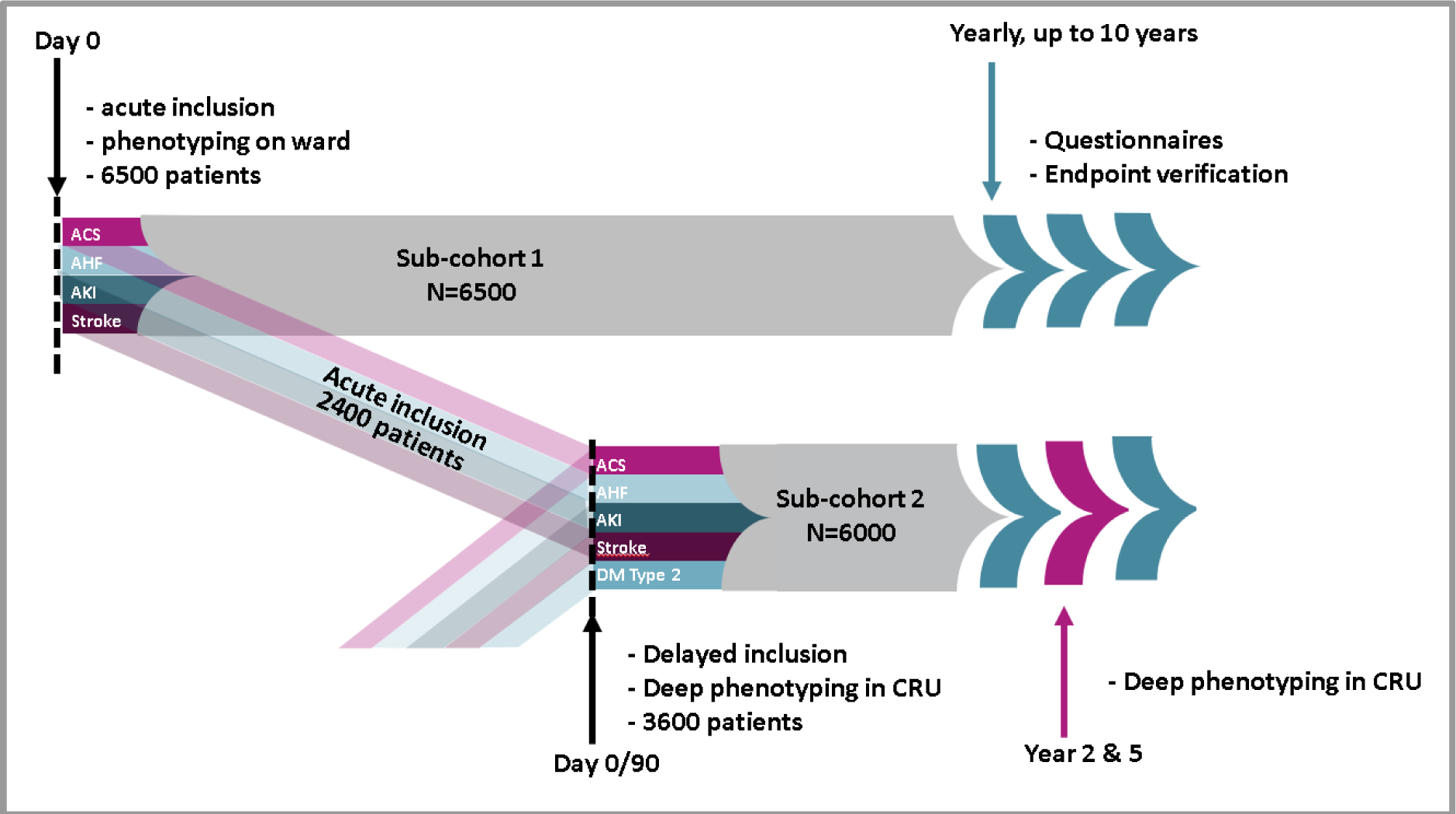
BeLOVE study design. provides an overview of the inclusion moments (acute and delayed) as well as the distinction between the two sub cohorts and their overlap. In this scenario, where all disease entities recruit optimal and a deep phenotyping participation of 37.5%, a grand total of 10.100 patients will need to be recruited. Both subcohorts will have yearly follow-up visits (telephone, postal, internet based) for verification of clinical endpoints as well as several other questionnaires (e.g. patient reported outcomes).Patients who underwent deep phenotyping will be reinvited for the same procedures 2, 5 and 10 years later.

The chronic subcohort (subcohort 2) includes up to 6000 patients with ACS, AS, AHF, or AKI 90 days after their event, as well as patients with type 2 diabetes and target-organ damage. These patients will undergo a 7-8 hour deep phenotyping program, including but not limited to the collection of biological samples (i.e. blood, urine, saliva, stool), and detailed questionnaires including Patient-Reported Outcome Measures (PROMs). Patients will also undergo a series of imaging procedures, including ultrasound, optical coherence tomography and magnetic resonance imaging. Follow-up of all patients is planned to continue for up to 10 years with annual telephone interviews, as well as visits to our clinical research unit 90 days, as well as 2, 5 and 10 years after the acute event or corresponding timeframes for T2DM. Patients are recruited from the Charité university medical center within the city of Berlin, Germany. The main phase of the study will start in November 2019 and is expected to recruit for up to 7 years..

### in- and exclusion criteria

In general, eligible for inclusion in the acute or chronic subcohort are patients with high cardiovascular risk, manifest through their clinical presentation (Table 1, Appendix 1). More specifically, this is defined as patients with one of the following 4 acute clinical events, being hospitalization for ACS (type 1,2, or 3 as defined by Fourth Universal Definition of Myocardial Infarction 2018), hospitalization for AHF (based on NYHA ≥II or overt clinical deterioration in combination with escalation or initiation of diuretic treatment), in-hospital stage 2 or stage 3 AKI lasting at least 72 hours (according to KDIGO criteria), hospitalization for AS (in accordance with the ASA guidelines). Additionally, patients who are being treated for T2DM with evidence of micro- or macrovascular damage at one of our specialized outpatient clinics for metabolic diseases are also eligible for inclusion into the chronic subcohort because of their equivalent high cardiovascular risk.

**Table 1:**
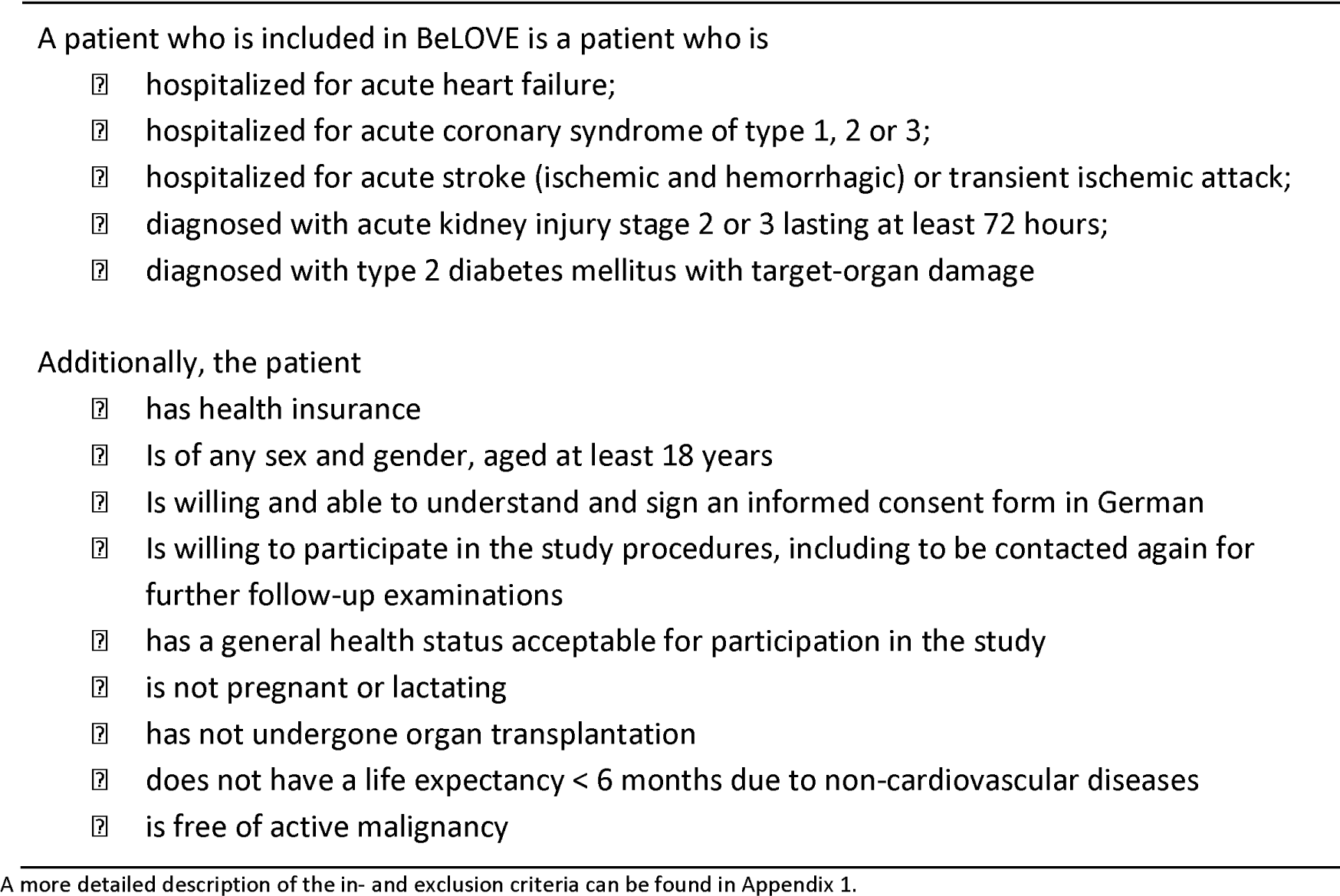
General in- and exclusion criteria.

Exclusion criteria are: <18 years at the time of study qualification, pregnancy or lactation, organ transplanted, lack of health insurance (health insurance is mandatory for all German residents). Additionally, the presence of active cancer (irrespective of treatment status), or a maximum life expectancy of six months or less due to non-cardiovascular disease as estimated by the study physician, patients who do not speak German, or cannot provide written informed consent, are similarly excluded from participation.

### Timing of inclusion

BeLOVE has two distinct time windows for inclusion. The first time window, referred to as “acute inclusion” marks the start of the acute subcohort. Acute inclusion is possible for patients with one of the four acute diseases (i.e. AS, ACS, AHF, and sustained AKI). These patients are contacted within 7 days after their qualifying event (goal day 1 - 2). This translates to a time window of up to 10 days after the start of the AKI episode to contact AKI patients, given the requirement of the AKI episode lasting at least 72 hours. Patients receive information regarding the goal and design of the study in both verbal and written form. After at least 12-24 hours, patients may sign the written informed consent. The data collection in the acute phase is limited in scope and comprises a limited set of questionnaires regarding pre-existing risk factors, agreement to use routine data produced during the hospital stay and subsequent visits to our hospital, as well as blood sampling.

The second moment of inclusion, referred to as “delayed inclusion”, is possible for patients with an acute qualifying event who were not able to provide consent during the acute event, as well as patients who deferred the decision to participate to a later moment. The second moment of inclusion is the only moment of inclusion for patients who qualify through the T2DM criterion. This second moment of inclusion falls together with the deep phenotyping visit to one of our three dedicated clinical research units (CRU) that marks the baseline measurement of our chronic subcohort. This subcohort thus comprises all patients from the delayed inclusion as well as the patient from the acute inclusion who take part in the CRU deep phenotyping program. These CRU visits are planned at day 90 after the qualifying event with a −7/+21 range (goal 90-97) after the initial qualifying acute event. All patients planning to participate in the CRU phenotyping program receive extra information regarding the next steps in the study protocol, as well as the possibility to start with the questionnaires.

Patients who declined any inclusion (acute or delayed) re-qualify if they experience a second qualifying event. If the second qualifying event is of a different phenotype as the initial event, the interval between the two qualifying events must be at least 7 days (10 days if AKI is the second qualifying event), irrespective of whether the patient was discharged or not. If the second event is the same as the initial qualifying event, the minimal interval is 60 days. Patients who qualify by the T2DM criteria but decline any participation are not approached for a second time unless they are treated in our hospital for one of the acute qualifying diseases.

## Data collection

### Acute phenotyping

Patients characterized by an acute event undergo a limited phenotyping procedure which includes fasting blood sampling, medical history and numerous self-assessed tablet based questionnaires, including PROMIS-29 (www.healthmeasures.net) [36] and EQ-5d-3L (www.euroqol.org)[37] and a series of disease-specific questionnaires (Table 2, Appendix 2). Paper questionnaires are available upon request. Cognition is evaluated by the Montreal Cognitive Assessment Instrument (MOCA) while mobility and dependency are assessed by the modified Rankin Scale (mRS). A 14-day continuous glucose monitor is applied to the patient’s upper arm which the patient can remove and send back to the CRU by mail. Blood samples, of which a portion is analyzed directly while another part is prepared for storage in our biobank, include serum, heparin, CPT, citrate, EDTA. Other acute biosampling also includes buffy coat for DNA extraction, dry blood, urine sampling in a subset of patients.

**Table 2:** General overview phenotyping program.

| Group of assessments | Time of assessment |  |  |
| --- | --- | --- | --- |
|  | Acute inclusion | At home questionnaire | CRU visit |
| Demographics and anthropometry | ? |  | ? |
| Health history, behavior and medication | ? | ? | ? |
| Clinical outcomes (endpoints) |  | ? | ? |
| Scales and scores for symptoms/activities of daily living | ? | ? | ? |
| Metabolic function |  | ? | ? |
| Cardiovascular function |  |  | ? |
| Imaging |  |  | ? |
| Physical function |  |  | ? |
| Somatosensation |  |  | ? |
| Electroencephalography (ECG) |  |  | ? |
| Cognitive function | ? | ? | ? |
| Disease specific quality of life (QoL) | ? | ? | ? |
| Health related QoL and Patient-Reported Outcomes | ? | ? | ? |
| Biosampling |  |  |  |
| · Blood sample LB (immediately analyzed) | ? |  | ? |
| · Blood sample BC (Immune phenotyping) | ? |  | ? |
| · Blood sample BB (storage) | ? |  | ? |
| · IPS-programming from blood cells | ? |  | ? |
| · Urine | ? |  | ? |
| · Stool |  |  | ? |
| · Saliva sample |  |  | ? |

A more detailed description of the phenotyping program, including the source of the data, as well as a more detailed overview of the timing, can be found in Appendix 2.

### CRU phenotyping

Following a 12-hour overnight fast, all patients are investigated at one of the three CRU sites starting at 8.00 a.m. During this visit, a series of questionnaires is completed, focussing on demographics, lifestyle, socioeconomic status, health history, reproductive history, food intake, and general health status, quality of life and patient reported outcomes. Further assessments include, but are not limited to clinical symptoms, anthropometry, metabolism, frailty and physical function, different domains within physical activity and neuromuscular and somatosensory function, as well as cognitive function (e.g. MOCA and the CANTAB Connect Test Battery). Other assessments include a 12-channel-electrocardiography (ECG), 64-channel dry electroencephalography (EEG), and calorimetry [38]. The patients are also requested to participate in the assessment that extends beyond the CRU visit itself, such as 24-hour Holter ECG, 7-day accelerometer and again a 14-day continuous glucose monitoring. Several blood samples are taken in a fasting state and include serum, heparin, CPT, citrate, EDTA (including aprotinin and FC mix) and Tempus™ for RNA extraction. Other bio-sampling includes, urine, saliva, and stool. These biosamples are, after pre-analytical processing and aliquotation, initially stored at −80 degrees or liquid nitrogen in our centers’ central biobank.

### Imaging

Imaging includes a standardized 2D-echocardiography (optional 3D measures) as well as the measurement of carotid intima-media thickness (IMT) by trained and certified study personnel and assessed in a dedicated blinded echo core-lab. Moreover, three-dimensional imaging of retinal vessels, retinal blood flow as well as neuronal architecture will be assessed by optical coherence tomography (OCT). Data from coronary OCT-angiography performed in selected ACS patients as part of their routine care interventions will also be collected. After the visit to the CRU, eligible patients are invited for a separate visit to one of our four facilities for magnetic resonance imaging (MRI) and MR-spectroscopy (MRS). MRI includes assessment of the brain to detect even small ischemic injuries or functional changes (e.g. (sub)clinical ischemic infarcts, white matter lesions, cerebral microbleeds) as well as cardiac MRI for the quantification of function, deformation and structure of the heart including quantification of fibrotic remodeling. Fat metabolism will be evaluated by abdominal fat imaging and MRS.

### Testing hemostasis: nutritional and exercise induced changes in biomarkers

One of the major research questions of the study is the relation of nutritional or exercise-induced changes of blood based biomarkers on the development of future cardiovascular events. The idea behind this is that inability to maintain or quickly restore homeostasis could be a strong marker of underlying cardiovascular risk.[39] To test this, patients are randomized (stratified on study site and qualifying disease) to either a nutritional challenge (substrate coping) or a physical challenge (substrate expenditure), both performed in fasting state. The nutritional challenge includes a standardized mixed meal test meal (20% protein / 25% fat / 55% carbohydrates; 500 kcal) after 12 hours of fasting. The physical challenge includes a cardiopulmonary exercise test (CPET) using a spiroergometry cycle device according to established SOP from the German Center for Cardiovascular Research. To monitor and quantify performance, oxygen saturation will be measured continuously while a few drops of blood - usually from the ear lobes-will be taken sporadically to determine oxygen and carbon dioxide content and metabolic variables. Patients with contraindications for one of the challenges are not randomized but can participate in the other challenge. Blood samples will be collected at time of maximal exercise load (only CEPT) and 120 minutes after the test.

### Incidental findings

The phenotyping program is expected to lead to a considerable rate of (incidental) findings with potential clinical relevance. Since the clinical interpretation of results and direct treatment decisions are neither desired nor feasible within in our study setting, prompt communication of incidental findings with potentially high and urgent clinical relevance to clinical teams is pivotal. The subsequent triage acknowledges the desire to keep the number of false positive as low as possible for both patient and the healthcare system. To handle these issues BeLOVE has established a standardized findings management. Based on ethical guidelines [40–42], evidence-based recommendations from specific medical guidelines, procedures of other studies[43,44] and our own experience from the pilot phase, an extensive standardized operating procedure was developed which defines specific results and their urgency to be communicated and acted on.

### Outcome ascertainment and adjudication of clinical outcomes

An extensive catalog with detailed definitions of all relevant clinical outcomes was developed. Vital status and clinical outcomes will be assessed yearly through a systematic search of the Charité’s electronic hospital information system. Additionally, participants will be interviewed by phone for the occurrence of new inpatient treatments, outpatient invasive procedures, and occurrence of new diseases. For participants not responding to calls information is requested through mail. If this fails, a formal request to the civil registration office will be carried out to clarify if the person has moved house or deceased, including the date of either of these events if applicable. In case a death occurred while the patient was in the hospital, medical records will be obtained. If the death occurred outside a medical facility the death certificate will be formally requested at public institutions.

A clinical event committee consisting of board-certified physicians of the responsible disciplines (cardiology, endocrinology, nephrology, and neurology) will adjudicate all clinical events (fatal and non-fatal) on the basis of written medical documents according to a standardized protocol. The committee will validate the information that is key to the diagnosis from the source data and subsequently classify the diagnosis according to a detailed pre-defined event definition. In case of uncertainty, a preassigned second committee, consisting of the PIs of the study, will make the final decision. This second committee will reach a conclusion based on the same source information as is used in the first evaluation. The results of the complete procedure are entered in an eCRF, which forms the basis for all later analysis involving adjudicated clinical events.

### Outcomes of interest

The main outcome of interest are Major Adverse Cardiac Events (MACE). MACE is a composite outcome consisting of cardiovascular mortality, non-fatal stroke, non-fatal myocardial infarction, or rehospitalization due to heart failure according to ESC-Guidelines [45]. Cardiovascular deaths are defined as sudden cardiac death, sudden death due to acute MI (MI type 3), death due to heart failure or cardiogenic shock, death due to stroke, or death due to other CV causes. Other secondary outcomes of interest are listed in the detailed endpoint catalog. Most important examples of these secondary endpoints are vascular disease recurrence, peripheral vascular events, diabetic microangiopathy and the diagnosis of incident diabetes, renal events, all-cause mortality, all other hospitalizations as well as all out- and inpatient treatments related to the BeLOVE findings management. Further outcomes measure the patient’s’ generic and disease specific Quality of Life (see Appendix 2 for a full overview).

## Study size, precision, and minimally detectable effect sizes

BeLOVE is an observational study with a plethora of research questions, each with their own idiosyncrasies when it comes to power or precision. A single or uniform calculation of the precision and minimal detectable effect sizes for all upcoming analyses is therefore not possible. Some considerations, however, can be mentioned, as they help convey the range of the expected precision that can be achieved within BeLOVE. This includes our focus on reporting of effect estimates and corresponding confidence intervals where possible and appropriate instead of reporting ‘statistical significance’, as is recommended by the STROBE guideline for the reporting of observational studies [46]. Still, traditional power statements work under the principles of frequentist statistics. Therefore, our detailed sample size justification as depicted in appendix 3 follows those principles while keeping the minimally detectable effect sizes, based on Cox proportional hazards regression. For example, with 6000 patients in BeLOVE chronic subcohort, an alpha of 0.05, an annual risk of MACE at 10% and correlation between covariables set at R2=0.3, we will be able to detect a hazard ratio of 1.15 with 80% power and 1.17 with 90% power per standard deviation increase of a biomarker. For binary exposures with 50% prevalence, these numbers are 1.31 and 1.37 respectively. The acute subcohort will have a slightly increased precision as the expected sample size is 6500. If we want to study a smaller subset of patients, e.g. n=2400 when comparing two patients groups, the minimally detectable effect sizes increase correspondingly, with 1.24 and 1.28 for the standardized effect and 1.54 and 1.65 for the binary exposure with 50% prevalence, respectively. Some have suggested to completely forego on the binary concept behind hypothesis testing in both frequentist and Bayesian tradition [47]. However, we believe that for some specific types of analyses, such as -omics analyses, hypothesis testing is and will be at the heart of the current analyses techniques in some fields. If used, appropriate analysis techniques will be applied to avoid unacceptable type I error rates, ranging from the traditional and conservative Bonferroni correction to or other concepts like false-discovery rate.

## Data-management and -storage

BeLOVE makes use of an open source electronic data entry and data management system (REDCap) [48]. Manually captured data (e.g. self-administered questionnaires, interview results, and results of bedside examinations) are collected web-based with a central electronic case report forms (eCRF) on a tablet. Data from medical devices will be captured through automated procedures for electronic data transfer to central data repositories. Similarly, measurements performed in biosamples which are not stored in the biobank will be processed through a central laboratory information management system (LabVantage) and transcribed to the REDCap eCRF. The repository for all laboratory data, including metadata, is located on a database server which is centrally managed. This management includes central execution of data validation procedures as well as query management. A master participant index, the management of pseudonyms and a central electronic informed consent management will be performed by a trusted third party separate from the main study database.

## Quality assurance and control measures

BeLOVE aims for the highest quality in data collection, analyses, and reporting. Therefore, a comprehensive quality assurance and control concept is key. Our QA and QC concept was developed and will be constantly be updated, by the central structures for internal and external quality management at the Clinical Research Units of the Berlin institute of Health / Charité. Our concept is in line with the more general published principles and guidelines for Good Clinical Practice (ICH-GCP), Good Laboratory Practices (GLP) and Good Epidemiological Practice (GEP). The implementation of standard operating procedures for all elements of data collection and a delegation log of responsibilities will help standardize our efforts. This includes the periodic calibration of data capturing devices to reduce measurement errors and batch effects. More importantly, the training, certification of all personnel involved in collection of data and biosamples, as well as the continuous testing of our data collection procedures will help to ensure high-quality data collection throughout the study. This is supported by data monitoring that to ensure that the rights and well-being of participants are protected, that the reported study data are accurate, complete, and verifiable, and that the conduct of the study is in compliance with the currently approved protocol/amendment(s), with GCP, and with the applicable regulatory requirements.

## Open science

Several rules and safeguards for open science are developed and will be kept in line with the developments in open science by our use- and access committee, as well as our publication committee. Further, BeLOVE publications should be made open access, preferably through publication in open access journals (or the open access option within subscription journals (golden route to open access), or alternatively by depositing the final accepted version (post-print, maximal 3 months after publication) in the institutional repository of the free university Berlin (https://refubium.fu-berlin.de/) or the Max Delbrueck Center (https://edoc.mdc-berlin.de/).

## Data access and -sharing

Aggregated data will be reported on our website, allowing researchers to see if the BeLOVE study could be a relevant data source for their projects. Every researcher interested in the data of BeLOVE may apply for data access through our use and access committee, as long as one member of the project team is part of the BIH research community. The use and access committee evaluates the merits and technical feasibility of the project proposal and assesses potential overlap with ongoing projects and analyses. We support the publication of a BeLOVE dataset/analyses file as supportive information to a BeLOVE publication.

## Ethics and registration

The BeLOVE study is performed in accordance with the General Data Protection Regulation of the European Union as well as with other pertinent legislation and directives. A data protection concept according to the data protection recommendations of the platform for technology and methods for networked medical research (TMF; www.tmf-ev.de) has been established and is advised by the TMF as well as the official data protection officer of the Charité. Study protocols were approved by the Charité’s Ethics Committee. The statements of the German Ethics Council on human biobanks as well as big data and genetics research, as well as other ethically relevant directives, will be observed. He pilot phase of BeLOVE is registered at German Clinical Trials Registry DRKS00016852. The main phase will be registered at clinicaltrials.gov before inclusion start.

## Experiences from the pilot phase

The design and organization of the BeLOVE study have been an iterative process. What is described in this paper is the result of a pilot study, which started in 2017 and continues on up to the date of this publication, that initially focussed on establishing (first 147 patients) and later optimizing the logistics of the data collection. Some design elements, such as the possibility for delayed inclusion have been added to the BeLOVE design for the main phase, which is planned to be initiated in November 2019. Before that moment, all patients included in BeLOVE had to undergo the acute phenotyping in order to be eligible for delayed phenotyping. The phenotyping program has also been modified and optimized, based on patient experience, logistics, and costs, mainly in the form of a reduction of the program, in the hope to achieve higher recruitment and adherence rates. Wherever possible, we also adopted the randomization procedure for the challenge, allowing people to participate in the meal challenge if they were allocated but unable to perform the ergometer test and vice versa. Implementation of the recruitment procedures was different for each disease entity, leading to a different recruitment rate and total included patients. Given the changes in the study protocol, data collected in the pilot phase of the BeLOVE study are not incorporated in the BeLOVE core dataset. However, for some future analyses, it might be possible to merge the core dataset and the pilot dataset in order to increase precision. A comprehensive plan to deal with potential sources of bias, batch effects, and missing data needs to be submitted to the use and access committee by BeLOVE researcher who wishes to use the data of the pilot phase in their analyses. This plan should at least describe that all results will be presented in a stratified fashion in an appendix of the main publication.

## Discussion

BeLOVE is among the first few studies to examine disease overarching risk factors and pathways in high cardiovascular risk patients “over time”. The intensified deep phenotyping in BeLOVE will allow to study the short and long-term fate of patients with a high cardiovascular risk. BeLOVE will thus enable profound insight in common mechanisms underlying a whole group of diseases constituting one of the most significant and challenging burdens for health internationally.

Our inclusion procedures embedded within BeLOVE will create two subcohorts who overlap but are not nested in one other (see also figure 1 and figure 2). The acute subcohort has, by design, only a limited phenotyping program, while the chronic subcohort is characterized by a more detailed phenotyping program. Even though both sub-cohorts focus on the long-term cardiovascular risk, this detailed phenotyping program make cross sectional analyses also very interesting as they might identify disease overarching mechanisms in specific subgroups of patients. The clinical relevance of these mechanisms, for example in terms of improving risk prediction, can then be tested in both sub-cohorts in which timing phenotyping links up with two distinct, and arguably equally relevant, moments in clinical practice.

**Figure 2:**
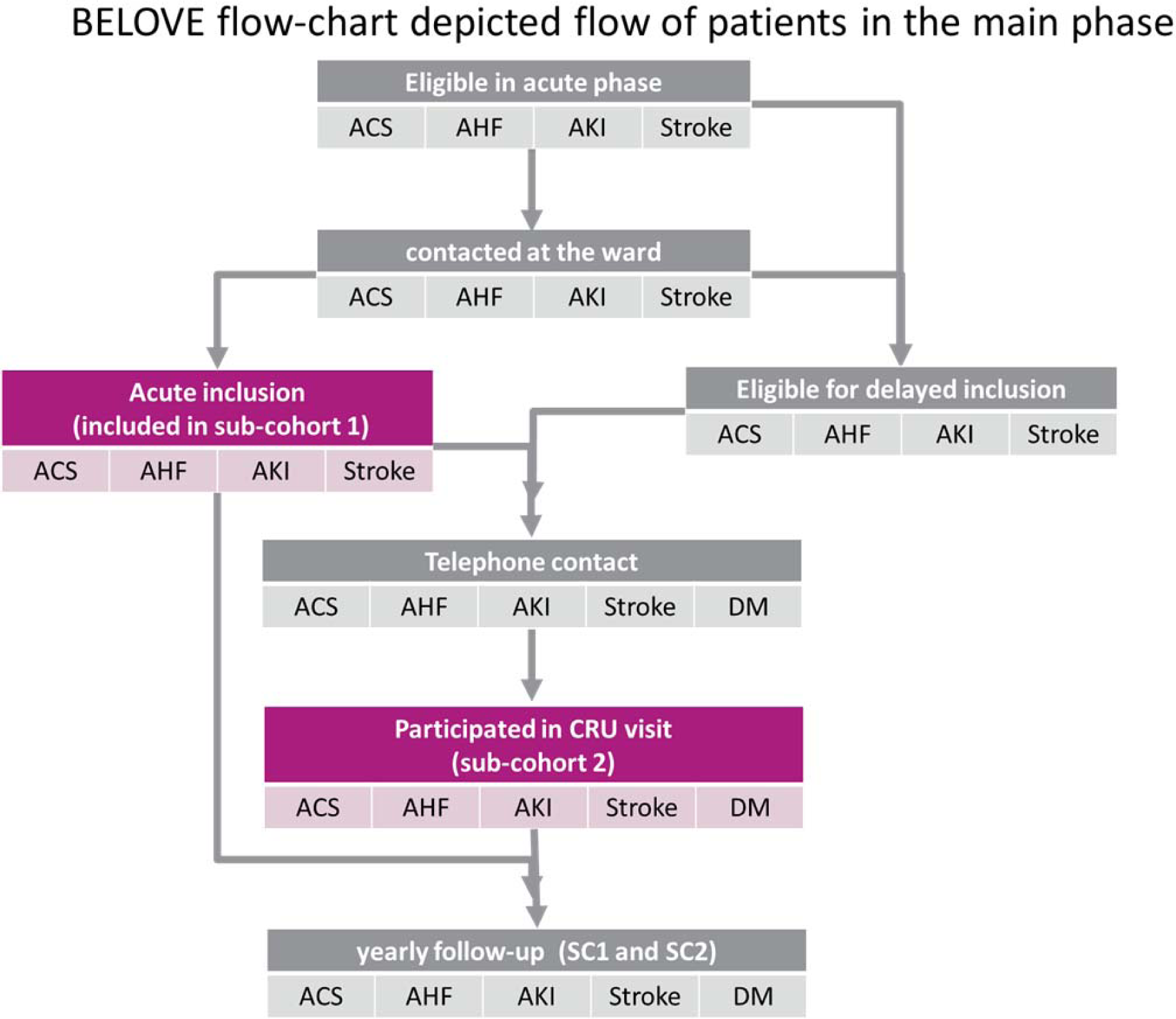
general flowchart of the BeLOVE, including subcohorts.

Even though the two moments of inclusion and phenotyping are chosen with the medical need in mind, not all research questions can be answered in either cohort. Issues, such as selection through self-selection and survival, could well lead to selection bias or at least limit the external validity of the results. Other sources of bias, including but not limited to, immortal time bias, prevalent user bias, and collider-stratification bias are not only possible but even likely threats to the internal validity - as is the case in any observational clinical study. An in-depth discussion of all the possible sources of bias is at this stage however not possible, as the relevance of these biases (direction and magnitude) will depend on the objective of the study (i.e. effect isolation in causal questions, or model performance in prediction and classification questions) as well as the subject matter of the specific biological mechanisms under study.

We believe that the methodological challenges in BeLOVE are to a large extent the same as any other clinical observational cohort. However, our two subcohorts approach shines a light on these shortcomings as we believe that some of the methodological disadvantages from one subcohort can mostly be counteracted by the advantages of the other subcohort. For example, the patients who are participating in the CRU visit are a selected subgroup due to patients declining or unable to participate (e.g. through death, worsened disease status, etc.). This selection is not unique to BeLOVE, but is, in fact, a limitation that all clinical studies with long phenotyping programs after the acute phase of the study need to to take into account. What is unique to BeLOVE is that with the combination both subcohorts, specifically the overlap between the two cohorts, we will be able to see and study the extent of these selection mechanisms at work. This will not only provide meaningful insights into the external validity of our results but could also add to the epidemiological literature on selection bias and its quantitative adjustment[49].

Due to limited resources, we have explicitly decided to not include some additional design elements in our study concept. Thus, BeLOVE does not include a population-based reference group which might have served as a reference point for cardiovascular morbidity and mortality but also to study the causes of the qualifying event using a case-control design. Given our primary aim of BeLOVE, we have decided to focus our resources on patient inclusion, as well as strengthening the long term follow-up procedures. Instead, we have opted for a cooperation with other cohorts for a reference group. Hence, we have standardized our data collection to a large extent by using common data elements (CDE) structure in order to make the comparison and combination of our data with other cohorts possible (www.nlm.nih.gov/cde). A good candidate for this is the German National Cohort or GNC study, a German population-based cohort study with 200K study participants of which enrollment started in 2014 [50]. An additional consideration was to add an additional blood sampling moment in the hyperacute phase, e.g. within 3-6 hours after arrival in the emergency room. Even though it would open up a plethora of research opportunities, it would also bring the moment of inclusion forward in time and thus reduce the number of patients who are willing and able to participate into the study. Of note, a delayed consent for non-interventional studies like BeLOVE or the concept of a broad general consent often cited in relation to biobank studies is currently not possible under current German law. The modular approach of BeLOVE makes it possible that smaller substudies - which could entail additional phenotyping within the existing BeLOVE framework but can also include additional study design elements - are embedded within BeLOVE. A complete overview of these modules - both current and former - can be found on the BeLOVE website.

We have to acknowledge the invaluable work that other cohort studies, old and current, did and do to examine factors that predict the outcome of cardiovascular disease patients. However, most of these studies focus on single disease entities, such as coronary heart disease or stroke. A study somewhat similar to BeLOVE is the Second Manifestations of ARTerial disease or SMART study [6,51]. Also using a disease overarching approach with a focus on crossover risk, this study initially planned to include a minimum of 1000 patients but currently included almost 10.000 CVD patients during 20 years of recruitment. In comparison to BeLOVE, SMART uses broader and more inclusion criteria leading to a heterogeneous study population, which range from risk factors (i.e. hypertension, hyperlipidemia and diabetes mellitus) to symptomatic arterial diseases (coronary artery disease, cerebrovascular disease, peripheral arterial obstructive disease or abdominal aortic aneurysm). Furthermore, this study does not have the same level of phenotyping and lacks the comparison between the acute phase and the post-acute phase, as well as the information on homeostasis maintenance gathered through our metabolic challenge. We believe that in BeLOVE, the combination of the deep phenotyping approach, the approach to investigate overarching risk factors and mechanisms across five different disease background, as well as the examination of risk factors in the acute phase as well as in the chronic phase does make our study stand out.

## Conclusion

BeLOVE provides a unique opportunity to study the short and long fate of patients with a high cardiovascular risk through state of the art deep phenotyping. With the same extensive standardized phenotyping procedures for all included patients that form the backbone of this study, we will be able to study disease overarching research questions and thus better understand crossover risk, as well as the similarities and differences between the different clinical phenotypes. Our unique design, with an acute and a chronic arm that overlap, allows us not only to study the short and the long-term but also allows us to understand and ameliorate potential biases. However, most importantly, the agility also provides us with the possibility to answer the research questions of the future, improving risk prediction and thereby individualized disease management.

## Data Availability

No data are reported in this manuscript.

## Acknowledgements

We would like to thank the Berlin Institute of Health, for their financial, operational and infrastructural support in the establishment of this interdisciplinary cohort. Additionally, the authors wish to thank all participants included in the BeLOVE pilot phase.

## Appendices

Appendix 1 - a detailed overview of the in- and exclusion criteria

Appendix 2 - a detailed overview of phenotyping program main phase

Appendix 3 - sample size justification and detailed power statement for the main phase

Appendix 4 - extensive overview of existing studies that touch some aspects of BeLOVE

Appendix 5 - BeLOVE contributors

